# Genotype and defects in microtubule-based motility correlate with clinical severity in *KIF1A* Associated Neurological Disorder

**DOI:** 10.1101/2020.07.27.20162974

**Authors:** Lia Boyle, Lu Rao, Simranpreet Kaur, Xiao Fan, Caroline Mebane, Laura Hamm, Andrew Thornton, Jared T Ahrendsen, Matthew P Anderson, John Christodoulou, Arne Gennerich, Yufeng Shen, Wendy K Chung

## Abstract

*KIF1A* Associated Neurological Disorder (KAND) encompasses a recently identified group of rare neurodegenerative conditions caused by variants in *KIF1A*, a member of the kinesin-3 family of microtubule (MT) motor proteins. Here we characterize the natural history of KAND in 117 individuals using a combination of caregiver or self-reported medical history, a standardized measure of adaptive behavior, clinical records, and neuropathology. We developed a heuristic severity score using a weighted sum of common symptoms to assess disease severity. Focusing on 100 individuals, we compared the average clinical severity score for each variant with *in silico* predictions of deleteriousness and location in the protein. We found increased severity is strongly associated with variants occurring in regions involved with ATP and MT-binding: the P-loop, switch I, and switch II. For a subset of identified variants, we generated recombinant mutant proteins which we used to assess transport *in vivo* by assessing neurite tip accumulation, and to assess MT binding, motor velocity, and processivity using total internal reflection fluorescence microscopy. We find all patient variants result in defects in transport, and describe three classes of protein dysfunction: reduced MT binding, reduced velocity and processivity, and increased non-motile rigor MT binding. The molecular rigor phenotype is consistently associated with the most severe clinical phenotype, while reduced binding is associated with milder clinical phenotypes. Our findings suggest the clinical phenotypic heterogeneity in KAND likely reflects and parallels diverse molecular phenotypes. We propose a new way to describe KAND subtypes to better capture the breadth of disease severity.

## Introduction

*KIF1A* Associated Neurological Disorder (KAND) encompasses a recently identified group of rare progressive neurodegenerative conditions caused by pathogenic variants in *KIF1A*^1-42^. Variants can be inherited dominantly or recessively, with *de novo* variants associated with the most severe phenotypes. A novel feature of KAND is the large number of disease-causing variants, predominantly missense variants within the motor domain of the protein. In the existing literature, 71 variants have been described to date, and here we describe 48 additional variants^2-34,37-39,41,43-45^. More than 30% of individuals with KAND have private variants, and there are likely many more variants that remain to be identified. KAND has a broad phenotypic spectrum, which can include spasticity, neurodevelopmental delay, intellectual disability, autism, microcephaly, progressive spastic paraplegia, autonomic and peripheral neuropathy, optic nerve atrophy, cerebral and cerebellar atrophy, and seizures^2-34,37-39,41,43-45^.

At the cellular level, KIF1A is a neuron-specific member of the kinesin-3 family of ATP-dependent microtubule (MT) molecular motor proteins. It predominately functions as a dimer and is responsible for anterograde transport of multiple cargos including synaptic vesicle precursors and dense core secretory vesicles on axonal and dendritic MTs^46,47^. The progression of kinesins along MTs requires ATP binding and hydrolysis, which cause conformational changes within the motor domain that power the forward motion of the motor^46^.

*KIF1A* has been associated with two different autosomal recessive conditions: hereditary spastic paraplegia (HSP, MIM:610357) and hereditary sensory and autonomic neuropathy (HSAN, MIM:614213)^2,3^. Individuals with biallelic missense variants in the motor domain were found to have HSP 30^3^, a condition consisting of peripheral nerve degeneration and severe distal sensory loss with distal motor involvement.

Heterozygous variants in *KIF1A* have been associated with NESCAV syndrome (Neurodegeneration and spasticity with or without cerebellar atrophy or cortical visual impairment, formerly mental retardation autosomal dominant 9) (MIM:614255), PEHO syndrome (Progressive encephalopathy with Edema, Hypsarrhythmia, and Optic atrophy) and with inherited cases of both simple and complicated HSP ^4-34,37-39,41,43-45^. More recently, individuals with clinically diagnosed Rett Syndrome were identified with *de novo KIF1A* variants in individuals who did not have variants in the genes that predominantly cause Rett syndrome (*MECP2, CDKL5* and *FOXG1)* ^35,40^.

Here we describe the natural history of KAND in 117 individuals with 68 different variants in *KIF1A*, provide detailed medical and neurodevelopmental phenotypes, and demonstrate a wide range of clinical severity with associated neuropathology on autopsy. We generated a KAND clinical severity score and correlate the clinical phenotype with genotype and molecular phenotype of the KIF1A motor.

## Subjects and Methods

### Cohort

This study was approved by the Institutional Review Board of Columbia University and informed consent was obtained for all participants. Individuals were referred by the family organization KIF1A.org. Clinical genetic test reports were reviewed to verify the diagnosis. Affected individuals or their parents or legal guardians completed a detailed structured medical history interview by phone and the Vineland Adaptive Behavior Scales, Third Edition (VABS-3) by computer. Clinical records and imaging data were reviewed to verify participant/caregiver reported history. Study data were managed using REDCap electronic data capture tools hosted at Columbia University Mailman School of Public Health^11,12^.

In two cases, postmortem brain specimens were collected through the Autism Brain Net (https://www.autismbrainnet.org/). Consent for autopsy was obtained from the decedent’s next of kin, which included provisions to save tissue for diagnostic and research purposes. The brain and spinal cord were removed and placed directly into 10% neutral buffered formalin fixed for 14 days before gross examination and sectioning. Formalin fixed paraffin embedded tissue was sectioned and stained with H&E and luxol fast blue (H&E/LFB) prior to histologic examination by two neuropathologists.

### Genetic Data Analysis

Individuals received clinical genetic testing from multiple clinical laboratories, with some individuals receiving whole exome sequencing and others panel gene testing. All *KIF1A* variants were re-interpreted centrally using ACMG guidelines^48^. Genetic reports were also reviewed for any additional potentially contributing genetic factors that might contribute to the phenotype (Table S1, S2).

Variants were annotated with their population allele frequencies in Exome Aggregation Consortium (ExAC) ^49^, Exome Sequencing Project (ESP; http://evs.gs.washington.edu/EVS/), 1000 Genomes Samples ^50^, and Genome Aggregation Database (gnomAD) ^49^. Computational scores from REVEL and CADD version 1.4 were collected using ANNOVAR version 2018Apr16. Biallelic predicted pathogenic variants that fit to an autosomal recessive inheritance pattern were identified. One individual was mosaic, and she was therefore excluded from the analyses to assess severity score, though her variant and her disease severity are shown in Figure 1.

**Figure 1.**
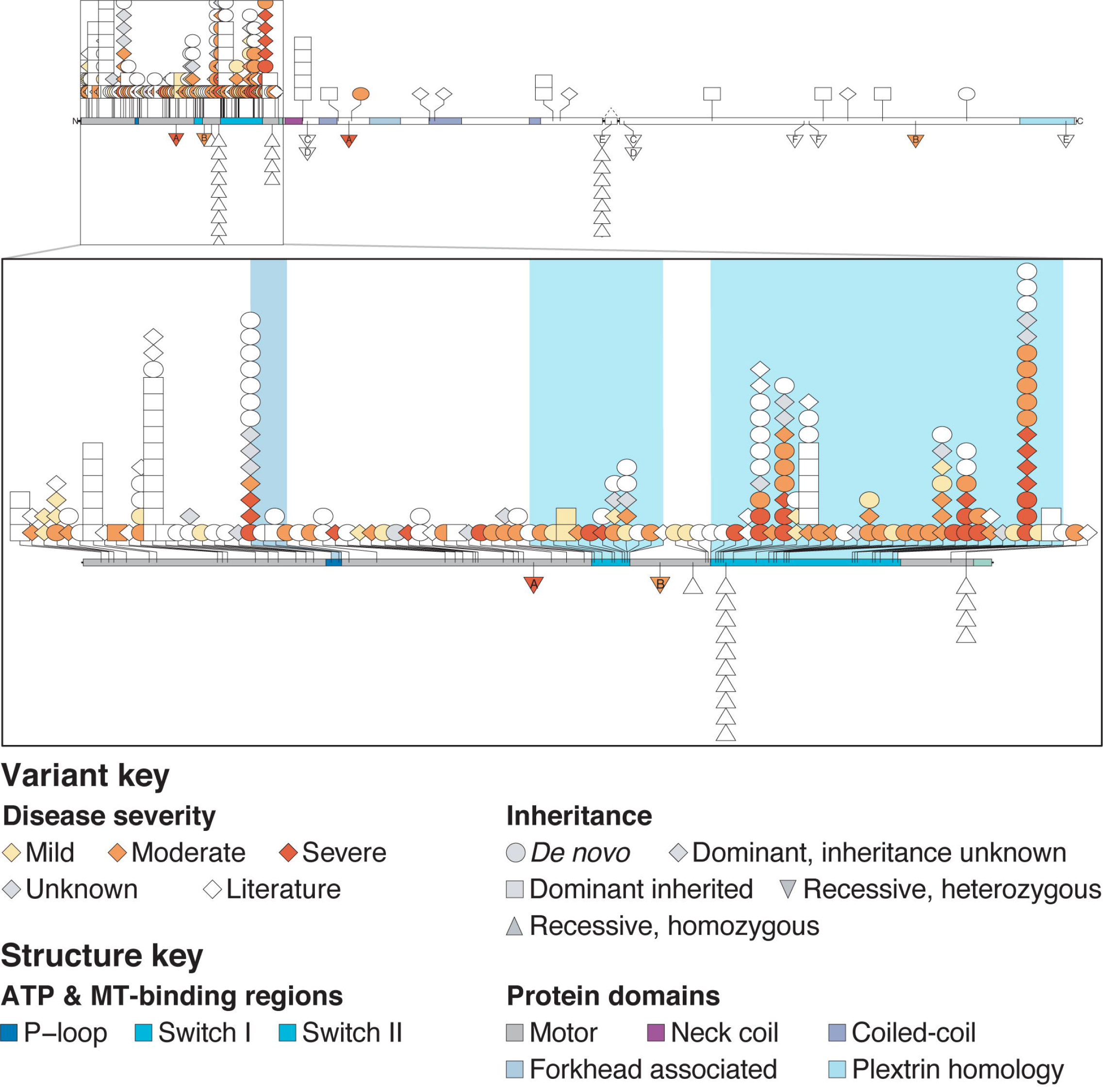
*KIF1A* variants by severity. Shapes representing 251 affected individuals are shown below, including 114 individuals from our cohort and 137 individuals previously reported in the literature. Disease severity is shown for 98 of these cases from our cohort. Four affected individuals are not incorporated as their variants are not mappable below: this includes 3 variants affecting splicing (2 in our cohort, 1 in the literature) and one larger deletion from the literature. On top is the full-length protein, and on the bottom in a zoom in of the motor domain. Each icon represents a unique individual. The shape indicates inheritance and color indicates disease severity. In both the full-length protein and the motor domain, monoallelic variants are shown above and biallelic variants are shown below. Compound heterozygous individuals are represented by two separate triangles, one at each variant: those triangles have letters to indicate variant pairing. ATP and MT-binding regions in the motor domain are indicated by color. Splicing information only indicated for the alternatively spliced exon 25b.

### Computational Methods

To summarize clinical severity, we used numerical values to represent the clinical features of brain atrophy, optic atrophy, seizure, microcephaly, developmental delay, spasticity, and hypotonia: yes (1), no (−1) and not sure (0); and converted VABS composite score (adaptive behavior composite, communication, daily living skills, socialization and motor skills) to 1 for VABS scores <70, 0 for VABS scores between 70 and 85, and −1 for scores >85. We designed a heuristic severity score by weighted sum of the medical history data. We set the weight to 2 for brain atrophy, seizure, and microcephaly, and the rest were given a weight of 1. We further performed a principal component analysis (PCA) in individuals who had both medical history and VABS data. We characterized the genetic variants using functional region of the protein, particularly ATP and MT-binding regions.

### Construct Generation

A recombinant *H. sapiens* KIF1A(1-393)-LZ-SNAPf-EGFP-6His construct containing the motor domain (amino acids 1-361), neck linker, neck coil domains, and a GCN4 leucine zipper (LZ) for stabilized dimerization was used for total internal reflection fluorescence (TIRF) microscopy. To generate this construct, a *R. norvegicus* KIF1A(1-393)-LZ-mScarlet construct obtained from Dr. Kassandra M. Ori-McKenney^51^ (UC Davis) was modified as follows: KIF1A(1-393) was amplified by PCR and inserted into a vector with a SNAPf-tag, GFP-tag, and a 6xHis tag at the C-terminus. *H. sapiens* KIF1A(1-393) is identical to *R. norvegicus*, except for Ile 359, which is Val in *H. sapiens*. The NEB Q5® site-directed mutagenesis kit (New England Biolabs Inc. #E0554S) was therefore used to induce the I359V mutation to generate the *H. sapiens* KIF1A(1-393)-LZ-SNAPf-EGFP-6His construct. This KIF1A construct was then used as template to generate the 8 mutants using the Q5 mutagenesis kit. All constructs were confirmed by sequencing.

For neurite tip accumulation assays, a KIF1A(1-393)-LZ-3xmCit construct tagged with three tandem mCitrine (mCit) fluorescent proteins at the C-terminus was obtained from Dr. Kristen Verhey (University of Michigan, USA)^52^. The sequences of all the constructs used in this project were analyzed using SnapGene Viewer software (version 4.0.2; https://www.snapgene.com/snapgene-viewer/; GSL Biotech). Primers flanking individual specific variation site were designed using web-based QuikChange Primer Design Program available online (www.agilent.com/genomics/qcpd) as per the software instructions. Patient specific variants were then introduced into the KIF1A(1-393)-3xmCit construct using QuikChange lightning mutagenesis kit (Agilent technologies #210519) according to the manufacturer’s instructions.

### Cell Lysate-Based Single-Molecule TIRF Assay

Expression of KIF1A constructs in *E. coli* bacteria for single-molecule TIRF imaging is described in detail in the Supplemental Material. Following the generation of the *E. coli* cell lysate and its clearance by centrifugation, the cleared lysate was used for the initial single-molecule TIRF microscopy studies to determine whether the mutant KIF1A motors are mobile or not. The assay was performed as described before^53^ except that a different assay buffer was used (see Supplemental Material). The lysate was diluted 100-fold in the assay buffer before the solution was flown into the slide chamber. The GFP-tag was used to track the motors. 600 frames were acquired for each video with an acquisition time of 200 ms per frame. Kymographs were generated using Fiji.

### Protein Purification

Further protein purification was performed as described in detail in the Supplemental Material. Following the incubation of the cleared *E. coli* cell lysate with an Ni-NTA resin, the resin was washed and then labeled with a SNAP-Cell® TMR-Star ligand. After another wash step, the protein was eluted with elution buffer and then flash frozen and stored at –80°C. Before further usage, an MT-binding and-release (MTBR) assay was performed as previously described^54^ with slight modifications (see Supplemental Material). The resulting protein solution was aliquoted and flash frozen as the “MTBR” fraction.

### Single-Molecule TIRF Assay With Purified Proteins

For the WT and mobile mutant motors, MTBR fractions were used for the single-molecule TIRF assay and the dilutions were adjusted to an appropriate density of motors on MTs. The TIRF assay was performed as described above for the studies with the lysate, except TMR was used to track the motors. Images were analyzed using a home-build MATLAB software, and graphs were generated using Prism.

### Neurite Tip Accumulation

SH-SY5Y (human neuroblastoma) cell-lines were grown in RPMI growth media (Sigma; Cat. no. R0883; with 3.7g/L NaHCO_3,_ 2.0g/L glucose and 0.3g/L L-glutamine) supplemented with 10% (vol/vol) FBS, 1X Pen-Strep at 37 °C in a humidified incubator and 5% CO_2_. The optimized density of SH-SY5Y cells (5*10^4^ cells per well) was plated on sterile glass coverslips coated with 3.3% rat collagen (Corning; Cat. no.354236) and 2% matrigel (Corning; Cat. no. 354248) in RPMI media without serum and antibiotics in a 12 well plate. After 24 hours, cells were transfected with the appropriate construct using Lipofectamine 2000 (Life Technologies; Cat. no. 11668019) according to the manufacturer’s instructions. Neuronal differentiation of the transfected cells was then induced using RPMI differentiation media (RPMI supplemented with 1% FBS, 1X Pen-Strep and 10µM retinoic acid (RA; Sigma; R2625)) that was refreshed every 24 hours for three days. Cells were then washed with warm 1X phosphate buffer saline (PBS; 137mM NaCl, 5.4mM KCl, 1.28mM NaH_2_PO_4_, 7mM Na_2_HPO_4_; pH 7.4), fixed in 4% (vol/vol) paraformaldehyde (PFA; Sigma; Cat. no. 158127) in PBS for 20 mins at room temperature before mounting on glass slides using Prolong Gold with DAPI (Thermo Fisher Scientific; Cat. no. P36935). Images were acquired using LSM780 fluorescence microscope with 40X objective (zoom factor =2). ImageJ was used to quantify the mean florescence intensity (MFI) of the expressed fluorescent protein along the neurite length and cell body of cells. Statistical analysis on the average MFI was performed using Mann Whitney in Prism software (GraphPad). All experiments were performed in triplicate and repeated at least two times prior to data analysis.

### Structure Rendering

All structures were rendered using Chimera^48^. The structure depicted is PDB 4UXP, *H. sapiens* KIF1A cryo-EM structure in the presence of AMP-PNP^55^. PDB 2HXF^56^ (cryo-EM structure of the *M. Musculus* MT-bound KIF1A motor domain) was used to assign the secondary structure to the residues.

## Results

### Summary of Clinical Phenotypes

A summary of the clinical, radiological, and neurodevelopmental features of individuals is shown in Table 1, with specific variant and severity information available in Table S1. We report 117 cases, of whom we have complete phenotypic data on 100 (Figure 1 and Table S1) While the majority of cases in our cohort had heterozygous *KIF1A* variants (115/117), we describe two biallelic individuals with asymptomatic carrier parents. In each biallelic instance there was one variant within the motor domain and one outside the motor domain. Among our heterozygous cases, the majority of *KIF1A* variants (64/115) were *de novo*, and 3 cases were inherited from an affected parent. Clinical information is not available for those affected parents. In some cases (48/115) parental testing results were not available. In a single instance, the affected individual was mosaic for the variant in blood.

**Table 1:**
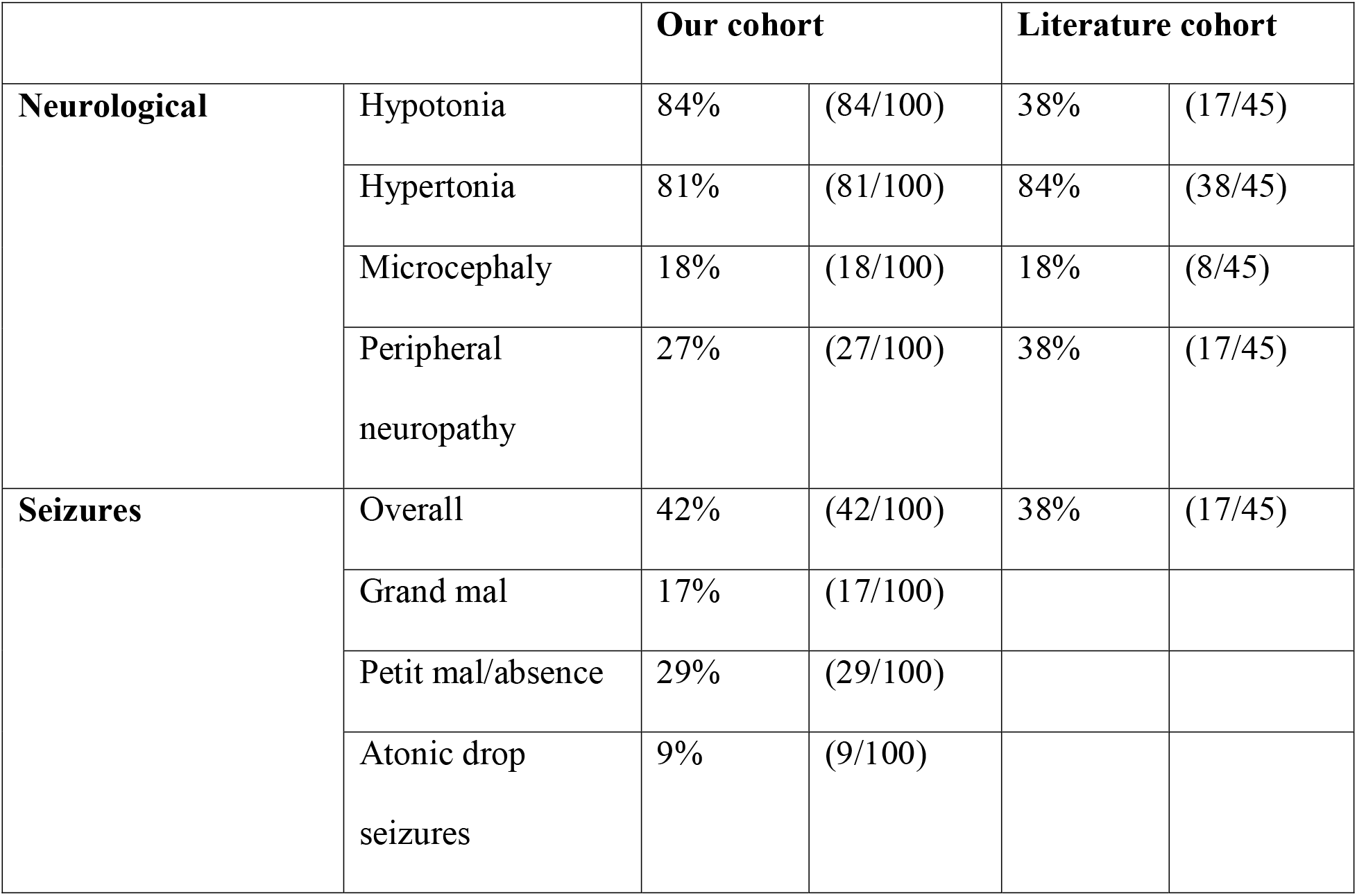
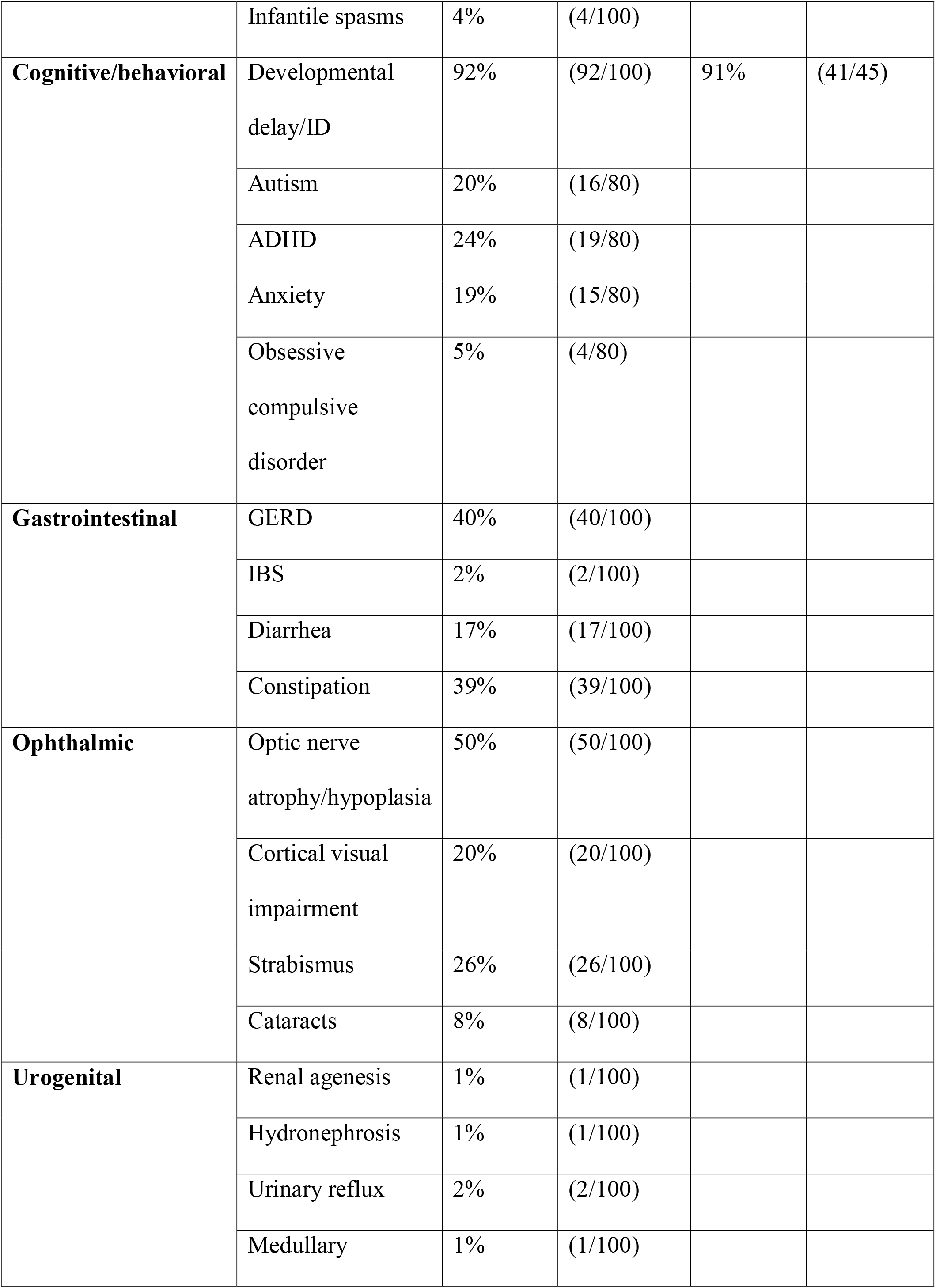
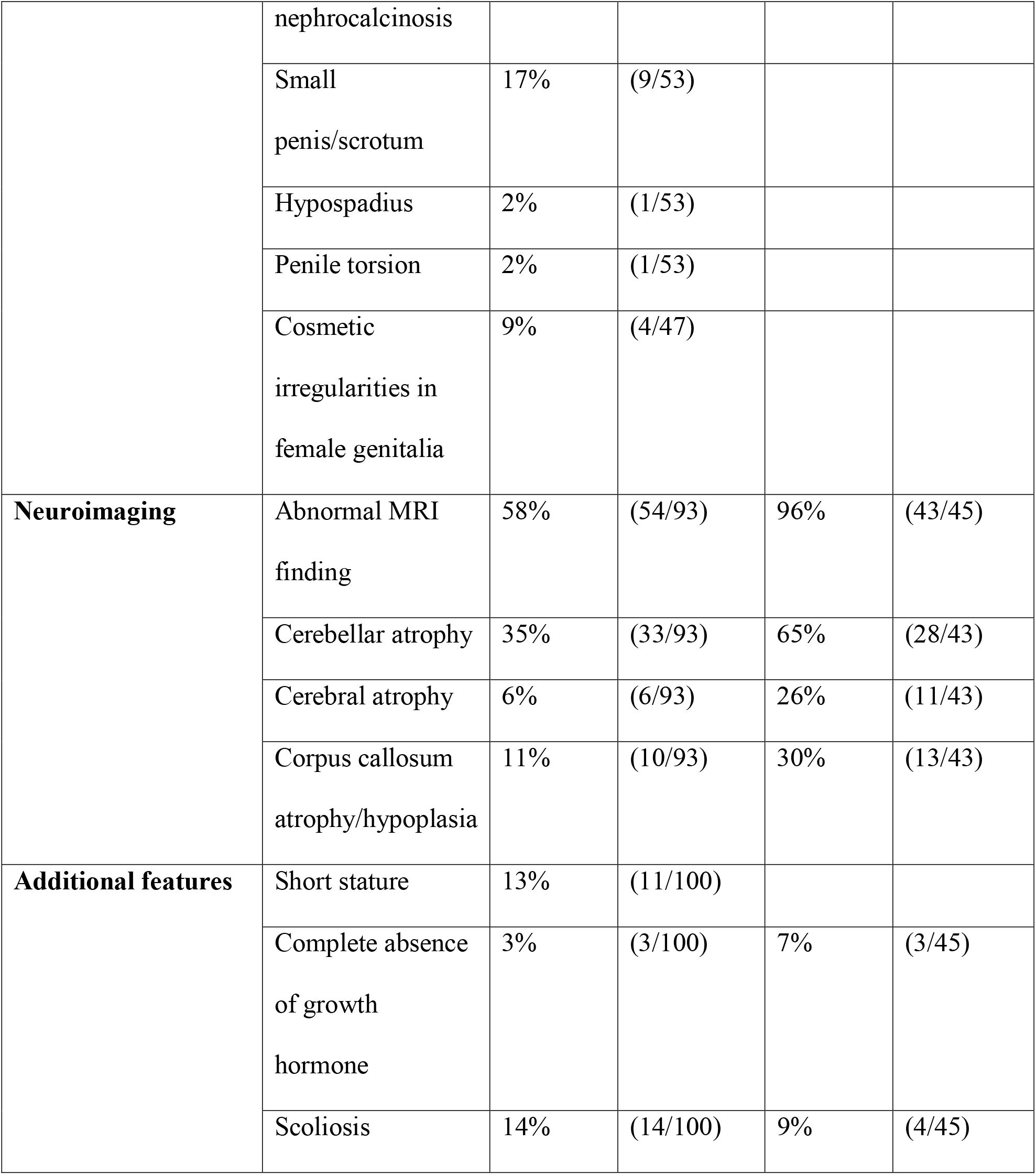
Phenotypic summary of KAND cases. Clinical phenotypes of individuals with variants in *KIF1A* are summarized by system. Table also includes summary data for 45 individuals from the literature with sufficient phenotypic information to allow for direct comparison with our cohort.

The most frequently observed symptoms in our cohort were hypotonia (84%, 84/100) and hypertonia or spasticity (81%, 81/100). Nearly all individuals had some degree of developmental delay and/or intellectual disability (92%, 92/100). Among those who had neuroimaging, the majority (58%, 54/93) had abnormal findings, most commonly cerebellar atrophy (35%, 33/93), abnormalities in the corpus callosum (11%, 10/93) and cerebral atrophy (6%, 6/93). Half of our cohort (50/100) had optic nerve atrophy or optic nerve hypoplasia, and there was a high prevalence of cortical visual impairment (20%, 20/100) and strabismus (26%, 26/100).

Developmentally, most individuals were globally delayed, with an average VABS-III Adaptive Behavior Composite score of 60.6 (population mean of 100, SD of 15) (Table 1, Table S2). Frequently reported neurobehavioral diagnoses included autism (20%, 16/80), attention deficit hyperactivity disorder (24%, 19/80), anxiety (19%, 15/80) and obsessive compulsive disorder (5%, 4/80). We also saw some features commonly seen in Rett syndrome, including stereotypic hand movements like wringing or clasping (21%, 17/80), small extremities (34% 27/90), cold extremities (60%, 48/80), bruxism (35%, 28/80), abnormal laughing spells (15%, 11/80), scoliosis (14%, 14/100) and high pain tolerance (65%, 52/80).

Seizures were common in our cohort, with 42% (42/100) reporting a history of seizures. Multiple seizure types were described, often in the same individual, with absence seizures as the most common in 29% (29/100) of the cohort overall and in 69% (29/42) of those with seizures.

We saw a number of features of dysautonomia. Just under half of individuals (46%, 37/80) experience difficulty with temperature regulation including sporadic fevers unrelated to illness. Gastrointestinal issues were common, with 40% (40/100) reporting gastroesophageal reflux disease (GERD), constipation (39%, 39/100) and diarrhea (17%, 17/100). Urinary retention and urinary incontinence were both reported. Many affected individuals experienced excessive salivation (26%, 21/80) or problems swallowing (29%, 23/80). Ten percent of the overall cohort, (10/100) required enteric nutritional support, in many cases due to the difficulty swallowing.

We saw additional features not previously described, including a small number of individuals with abnormalities in the urogenital system. Irregularities in external genitalia were seen in both males and females (10/53 males, 4/47 females) and a few individuals had structural renal issues including renal agenesis, hydronephrosis, medullary nephrocalcinosis, and urinary reflux. Another feature not previously reported in KAND was short stature, which we observed in 11% (11/100) of our cohort. In three of those individuals, complete growth hormone deficiency was noted, which has been previously observed^6,31,35^.

The condition is neurodegenerative, though the nature and course of decline varied by individual. Spasticity was progressive, and a number of individuals required surgical intervention: 15% (15/100) received tendon lengthening procedures. By the time they reached their 20s, many individuals were largely wheelchair dependent. When present, optic nerve atrophy also appeared to be progressive in most cases. While the loss of vision and of motor skills limited an individual’s ability to participate in the activities of daily living over time, we largely did not see any cognitive regression.

### Neuropathological Findings

At autopsy of a 13 year old female with a c.296C>T (p.Thr99Met) variant, the cerebellum was severely atrophic, most prominently in the midline, with relatively normal gyral patterning and development of the cerebrum (Figure 2A-F). Microscopic examination revealed severe cerebellar atrophy most prominent in the superior cerebellar vermis including a thin molecular layer, a depletion of Purkinje and internal granule cells, and Bergmann gliosis, and white matter pallor. Rare axonal spheroids were observed. The dentate nucleus as well as the inferior olivary nucleus of the medulla showed marked loss of neurons and severe reactive gliosis. Residual neurons in these areas were hypereosinophilic with pyknotic nuclei. Autopsy was also performed for a second patient with a *de novo* c.296C>T (p.Thr99Met) variant, a 5 year 9 month old female, and features were similar across both cases (Figure 2G-I).

**Figure 2.**
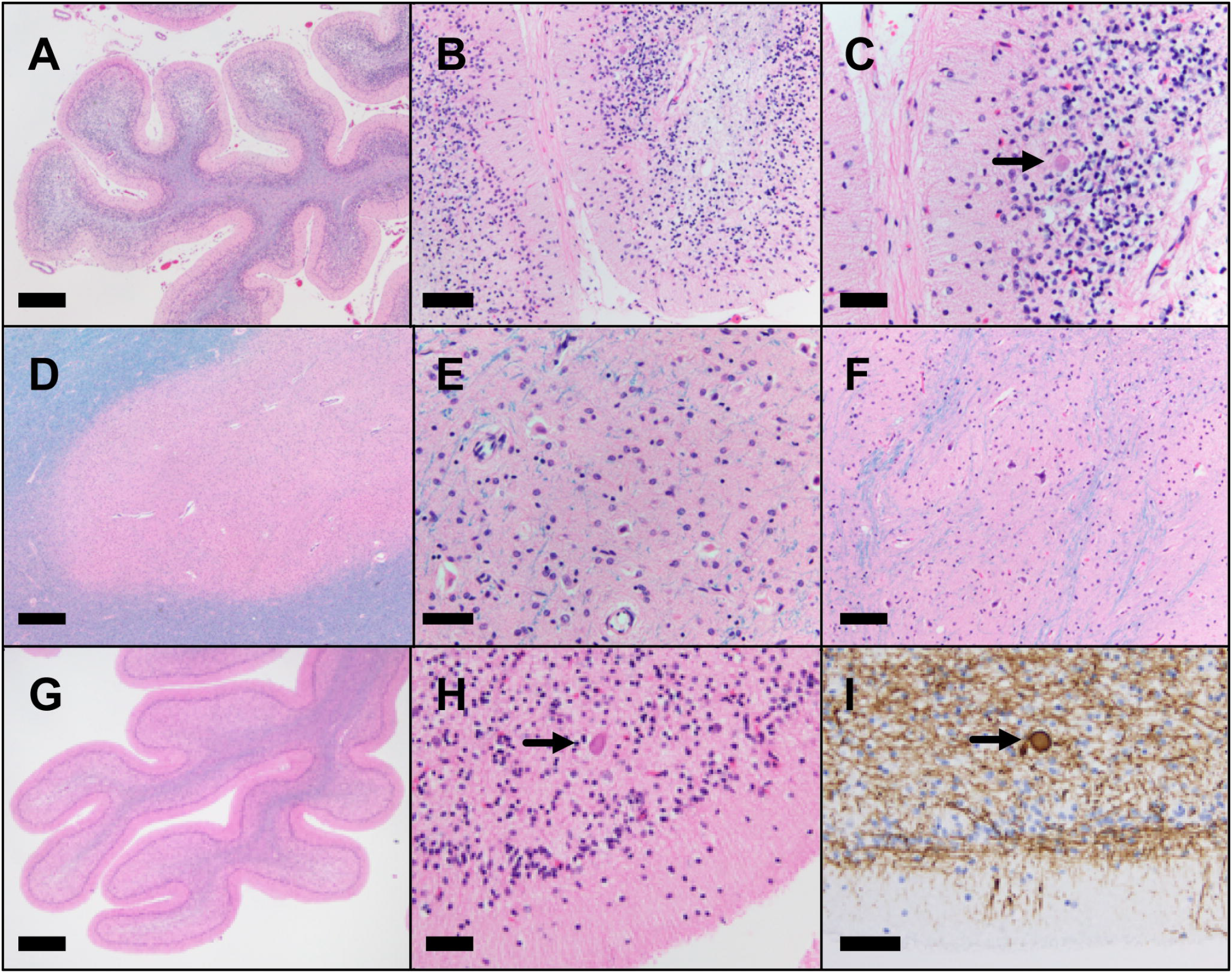
Neuropathological changes in two individuals with c.296C>T (p.Thr99Met) *KIF1A* variant. (A-F) Microscopic findings from a 13-year-old female (A-F) and a 6 year old female (G-I). Microscopic examination of H&E/luxol fast blue stained tissue sections reveal severely atrophic cerebellar folia (A, G) with thinned molecular layer, depletion of Purkinje cells, depletion of internal granule cells, Bergmann gliosis and pallor of the white matter (B,C,H). These changes are most severe in the superior aspect of the cerebellar vermis but are present in varying degrees throughout the cerebellum. Rare axonal spheroids are present (C, H, arrow) highlighted by neurofilament immunostain (I). The dentate nucleus (D,E) as well as the inferior olivary nucleus of the medulla (F) show marked loss of neurons and severe reactive gliosis. Residual neurons in these areas are hypereosinophilic with pyknotic nuclei. Scale bars: 1 mm (20x magnification) A, D, G; 200µm (100x magnification) B, F; 100µm (200x magnification) C, E, H, I.

### Computational results

Our heuristic severity score approximated a normal distribution across all cases (Figure 3A). Using all available medical history and VABS data, we limited the analyses to age, sex, and common symptoms (>15% prevalence in the cohort) with <10% missing data. Principal component analysis (PCA) showed principal component 1 (PC1) was attributed mostly to the same symptoms used in the heuristic severity score as well as the VABS scores. PC2 is correlated with the age of disease onset. Earlier-onset cases were more likely to develop microcephaly and later-onset cases more often underwent musculoskeletal surgeries. The first three PCs explained 13.5%, 7.8%, and 5.8% respectively of the variance in the data. Additionally, we found that the heuristic severity score was highly correlated (R=0.84) with PC1 (Figure 3B). Since PCA is an unsupervised analysis, this high correlation supported the conclusion that the heuristic score is a good overall measurement of KAND severity.

**Figure 3.**
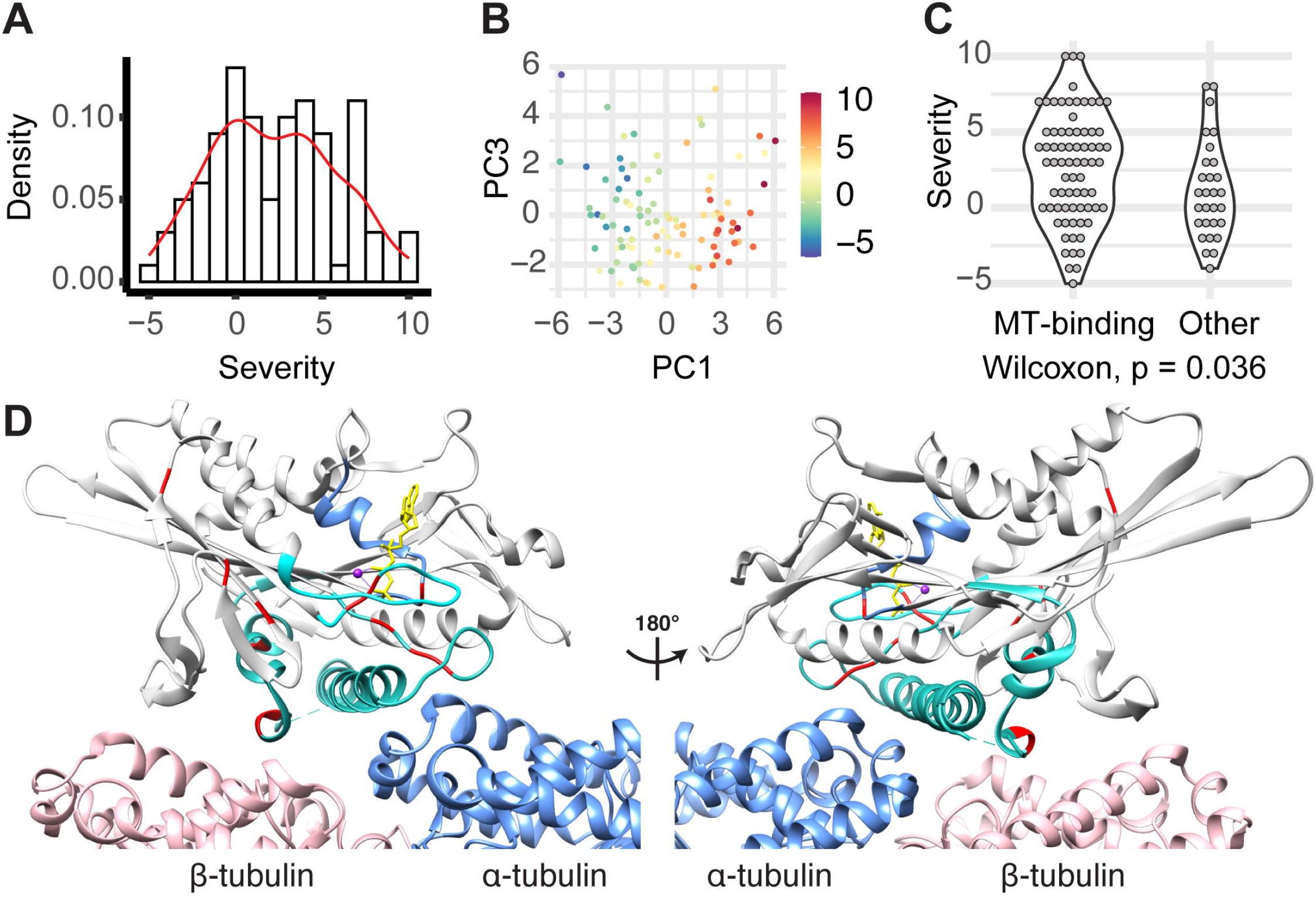
Clinical severity of 100 KAND cases. (A) Distribution of heuristic severity score. (B) Scatter plot of principle components 1 and 3 color-coded by medical history derived severity scores. (C) Violin plot of medical history derived severity scores for individuals carrying mutations in or outside of ATP or MT binding regions. (D) Structure of the MT-bound KIF1A motor domain with severe residues and functional regions involved in ATP and MT binding highlighted (PDB: 4UXP, *H. sapiens* KIF1A CryoEM structure in the presence of AMP-PNP). Light gray: KIF1A: light gray; Blue: α-tubulin; Pink: β-tubulin. Yellow: AMP-PNP; Red: residues mutated in most severe variants (p.Thr99, p.Glu148, p.Leu157, p.Val186, p.Ser214, p.Ser215, p.Gly251, p.Glu253, p.Arg254, p.Tyr306, p.Arg307, p.Arg316). The P-loop, Switch I and Switch II are shown in light blue, cyan, and teal respectively.

We hypothesized that severe cases have genetic variants with similar characteristics. We did not observe significant correlation between severity score and prediction score for variant deleteriousness such as REVEL and CADD, but all CADD scores were larger than 20 in this dataset. More importantly, the severity score was strongly associated with location in regions involved with ATP and MT-binding: the P-loop, switch I and switch II (Figure 3D). We found that 23 out of 28 (82%) of the variants seen in the most severe KAND cases (score ≥5) were located in the ATP and MT-binding regions, whereas only 14 out 24 (58.3%) of the least severe cases (score <0) had variants in these same regions (odds ratio = 1.4, p-value=0.003). Additionally, cases with variants in the ATP and MT-binding regions have significantly higher severity scores (p=0.036; delta mean=1.6) than the ones with variants outside the ATP and MT-binding regions (Figure 3C).

### Effect of KAND Variants on Neurite Tip Accumulation

To assess the effects of KAND variants on overall motor function of KIF1A, we expressed wild type (WT) and mutant KIF1A (1-393) tagged with three tandem copies of fluorescent monomeric citrine (mCit) in retinoic acid differentiated neuronal SH-SY5Y cells and measured the mean fluorescent intensity (MFI) at the cell body and along the neurite length. Consistent with previous work from our group and others, WT KIF1A accumulated at the distal tip of growing neurites in differentiated SH-SY5Y cells^35,52^ (Figure 4). All KIF1A variants tested showed significant abilities to accumulate along the neurites of differentiated cells.

**Figure 4.**
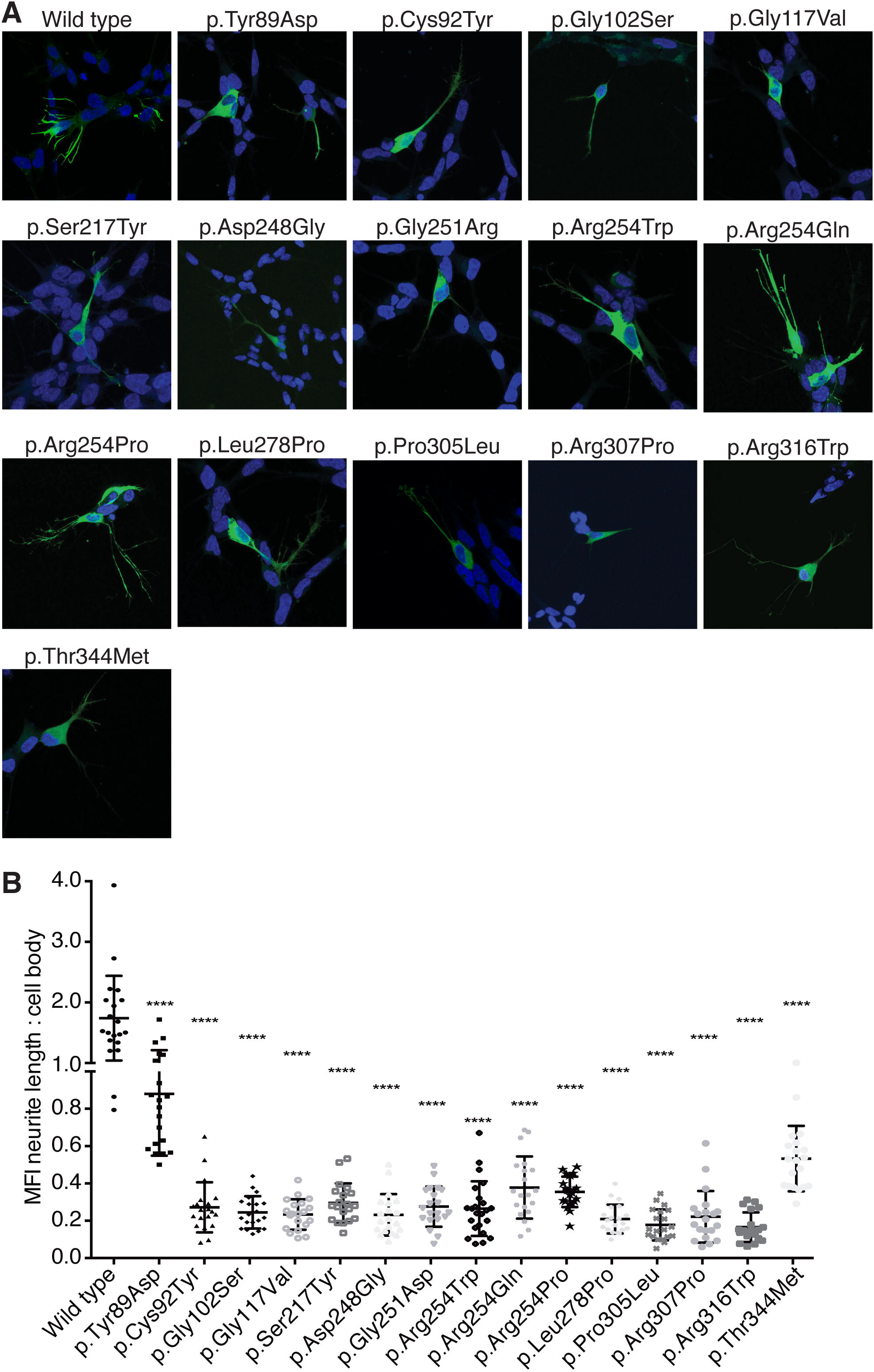
Neurite tip accumulation in differentiated SH-SY5Y cells for KIF1A variants seen in KAND individuals. (A) Rat KIF1A (1-393) tagged with 3xmCit tag (green) accumulated in the distal tips of differentiated SH-SY5Y cells in case of WT, but was localized closer to the cell body in cells transfected with KIF1A constructs containing individual variants. (B) A significant reduction in the mean florescence intensity (MFI) along the neurite length versus the cell body was observed in the 15 individual variants tested. Data were analyzed from three independent experiments using a Mann-Whitney test (n=20 for each experiment) and is plotted as mean ± standard deviation. **** represents p<0.0001 versus WT.

### Effect of KAND Variants on Velocity and Processivity

To determine the effects of KAND variants on the velocity and processivity (the ability to take multiple steps before dissociating) of KIF1A, we employed TIRF microscopy and a tail-truncated, constitutively active KIF1A (*H. sapiens* aa 1-393) with a C-terminal leucine zipper (LZ) for stabilized dimerization as well as a SNAP-tag and a GFP tag. WT KIF1A and mutant motors were expressed in *E. coli* bacteria. Our recent work has shown that the velocity and force generation properties of KIF1A expressed in *E. coli* bacteria and mammalian cells are statistically indistinguishable^57^. A similar construct has also been used in other studies^51,58,59^.

Eight variants were selected and generated by mutagenesis. The positions of the mutations are illustrated in Figure 5. The mutants c.38G>A (p.Arg13His), c.296C>T (p.Thr99Met), c.751G>A (p.Gly251Arg), c.757G>A (p.Glu253Lys), and c.760C>T (p.Arg254Trp) are at or close to the ATP-binding site, while c.821C>T (p.Ser274Leu), c.914C>T (p.Pro305Leu), and c.946C>T (p.Arg316Trp) are near by the MT binding site specifically at the β-tubulin interaction interface (Figure 5A). For an initial assessment of the effects of the mutations, we used the *E. coli* cell lysate with the expressed *KIF1A* mutants without further purification and directly performed single-molecule TIRF microscopy studies. These initial experiments revealed that while the c.760 C>T (p.Arg254Trp), c.821 C>T (p.Ser274Leu), c.914C>T (p.Pro305Leu), and c.946C>T (p.Arg316Trp) mutants retained their ability to move processively, they appeared to have reduced run lengths (average distance traveled) (Figure 5B). In contrast, the c.38G>A (p.Arg13His), c.296C>T (p.Thr99Met), c.751G>A (p.Gly251Arg), and c.757G>A (p.Glu253Lys) mutants were completely unable to move (Figure 5B). We found c.296C>T (p.Thr99Met), c.751G>A (p.Gly251Arg), and c.757G>A (p.Glu253Lys) attach to MTs in rigor while p.Arg13His lost its ability to bind to MTs (Figure 5B).

**Figure 5.**
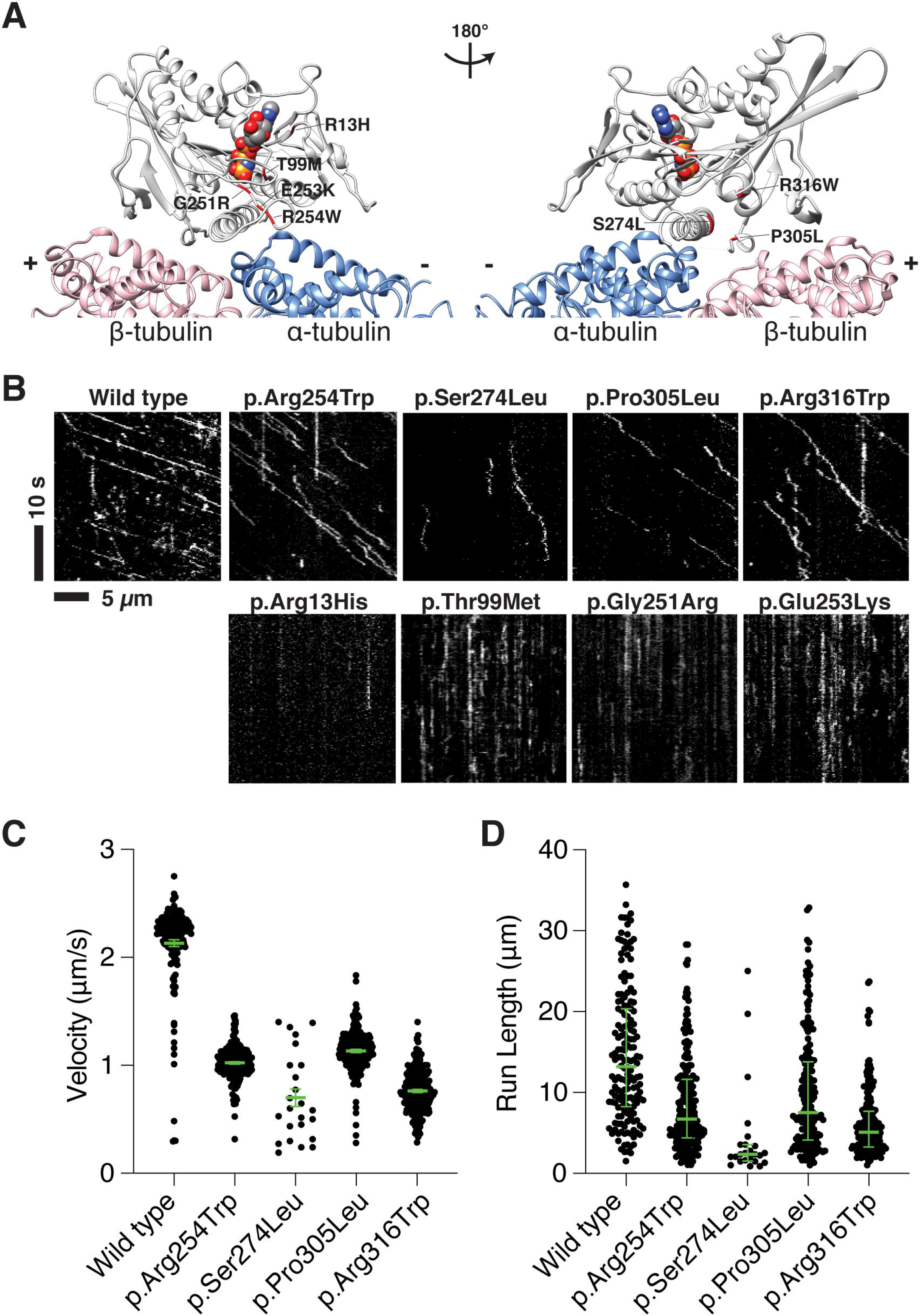
Sites of KIF1A mutations and motility analyses of dimeric KIF1A mutants. (A) Structure of the MT-bound KIF1A motor domain with the sites of mutated residues used for TIRF microscopy. Light gray: KIF1A; Blue: α-tubulin; Pink: β-tubulin. Van der Walls sphere represents AMP-PNP; red residues represent the mutation site (PDB: 4UXP, *H. sapiens* KIF1A CryoEM structure in the presence of AMP-PNP). p.Arg13His, p.Thr99Met, p.Gly251Arg, p.Glu253Lys, and p.Arg254Trp are at or close to the ATP binding site of KIF1A (left). p.Ser274Leu, p.Pro305Leu, and p.Arg316Trp are close to the β-tubulin binding interface (right). (B) Kymographs of KIF1A WT and mobile (top) and non-motile (bottom) mutant motors. Diagonal lines in the kymograph represent KIF1A molecules that are moving over time. The depicted scale bars are the same for all kymographs shown in this figure. (C) Velocities of KIF1A WT and the mobile mutants. The green bars indicate the mean with SEM. Compared to WT, all the mutants have reduced velocities. The WT has an average velocity of 2.1 ± 0.03 µm/s (mean ± SEM) (n = 162), while p.Arg254Trp, p.Ser274Leu, p.Pro305Leu, and p.Arg316Trp move at 1.0 ± 0.01 µm/s (n = 214), 0.7 ± 0.08 µm/s (n = 24), 1.1 ± 0.01 µm/s (n = 181), and 0.8 ± 0.01 µm/s (n = 210), respectively. (D) Run lengths of KIF1A WT and mutant motors. The green bars indicate the median with quartile. All the mutants have reduced run lengths. The WT’s processivity has a median value of 13.2 [8.3, 20.4] µm, while p.Arg254Trp, p.Ser274Leu, p.Pro305Leu, and p.Arg316Trp have run lengths of 6.7 [4.4, 11.6] µm, 2.3 [1.5, 3.5] µm, 7.5 [4.1, 13.8] µm, and 5.1 [3.2, 7.7] µm, respectively.

To further quantify the motility behaviors of the motile mutant motors, c.760 C>T (p.Arg254Trp), c.821C>T (p.Ser274Leu), c.914C>T (p.Pro305Leu), and c.946C>T (p.Arg316Trp) were purified via Ni-NTA affinity purification and labeled with a SNAP-tag TMR ligand, followed by MT-binding and release affinity purification^60^. All four mutants exhibited reduced velocities. While WT KIF1A has an average velocity of 2.10 ± 0.03 µm/s (mean ± SEM), c.760 C>T (p.Arg254Trp), c.821C>T (p.Ser274Leu), c.914C>T (p.Pro305Leu), and c.946C>T (p.Arg316Trp) move at 1.0 ± 0.01 µm/s, 0.7 ± 0.08 µm/s, 1.1 ± 0.01 µm/s, and 0.8 ± 0.01 µm/s, respectively (Figure 5C). In addition, all four mutants exhibited reduced processivities. While WT KIF1A is processive with a run length of 13.2 µm (median value), c.760 C>T (p.Arg254Trp), c.821C>T (p.Ser274Leu), c.914C>T (p.Pro305Leu), and c.946C>T (p.Arg316Trp) exhibited run lengths of 6.7 µm, 2.3 µm, 7.5 µm, and 5.1 µm, respectively (Figure 5D).

## Discussion

We describe the largest number of KAND individuals to date, including 48 novel missense variants, many of which were observed in single individuals. This suggests that we have not yet observed the full allelic spectrum in KAND. *KIF1A* is highly conserved. *KIF1A* has both fewer loss-of-function variants (o/e=0.07) and fewer missense variants than would be expected (0.57)^61^. We observed both dominant and recessive inheritance patterns, with the majority of KAND cases seen with heterozygous missense variants. Predicted loss-of-function variants were associated with milder disease than missense alleles.

The majority of individuals diagnosed with *KIF1A*-associated HSAN had biallelic loss-of-function variants in the alternatively spliced exon 25b, though one individual had, in trans, one loss-of-function variant in exon 25b and a second loss-of-function in exon 45^2^. In all families with recessive *KIF1A*-associated HSAN, individuals heterozygous for a single loss-of-function variant were asymptomatic. There were notable differences between the individuals with *KIF1A*-associated HSAN who had homozygous variants in exon 25b and the single individual who was heterozygous for variants both in exon 25b and 45. All the homozygous individuals had disease onset in early adolescence (mean 9.7 years, SD 3.0 years) in contrast with the congenital disease seen in the compound heterozygous individual. Similarly, none of the homozygous individuals had any reported cognitive issues or issues with gait, while the compound heterozygous individual had delayed speech development with poor articulation and an IQ of 80 as well as short stature and difficulties with gait/ambulation requiring use of a wheelchair^2^. However, there was only one individual with HSAN who had a loss-of-function allele outside of exon 25b, so replication is required.

Heterozygous loss-of-function variants including both premature termination and frameshift variants in *KIF1A* were associated with HSP. These individuals, some in their 60s at the time of report, all had normal cognition, normal MRIs, and no history of the visual problems or seizures common in our cohort^32^. If we hypothesize these loss-of-function variants result in nonsense mediated decay of the mutant transcript, this suggests that up to a 50% reduction in protein can potentially be tolerated with minimal clinical symptoms for many decades.

The clinical disease severity associated with missense variants in our study was much worse than any of the loss-of-function alleles, suggesting a dominant negative disease mechanism. Since KIF1A predominantly functions as a dimer, if the mutant and normal proteins dimerize to a similar degree, heterozygous individuals should only have 25% of the dimeric protein that is normal. Variants with decreased or no MT binding, like p.Arg13His and p.Ser274Leu (Figure S2), are associated with relatively milder disease, perhaps because mutant protein homodimers are unable to bind to MTs, and therefore cannot interfere globally with MT traffic, but only lead to decreased transport of KIF1A cargo. In contrast, variants including p.Gly251Arg and p.Glu253Lys, which bind tightly to the MT but are unable to engage in processive motion, were associated with the most severe disease. These rigor mutants likely lead to the most severe impact on all traffic along MTs, and may act as roadblocks resulting in an overall deficiency of cargo transport along the associated MT^62^. For alleles, like p.Arg254Trp, p.Pro305Leu, and p.Arg316Trp, where the mutant proteins bind to MTs but show reduced processivities and velocities (Figure 5C&D), *in vivo* these kinesins may affect MT transport globally and could interfere with all traffic along MTs.

While a previous study suggested that the p.Arg254Trp and p.Arg316Trp mutants were incapable of binding to and moving along MTs, we found that these mutants were processive (the published results on the p.Arg13His, p.Thr99Met and p.Glu253Lys mutants agree with our findings) ^59^. The reason for this discrepancy is unclear.

Consistent with previous reports, the majority of disease-associated *KIF1A* variants we report are located in the protein’s motor domain. Using our clinical severity score, we observed that the most severe phenotypes were associated with variants clustered near the ATP and MT binding sites (Figure 5A). This includes the p.Thr99Met, p.Gly251Arg and p.Glu253Lys variants whose rigor phenotype likely represents defects in KIF1A’s capacity to hydrolyze ATP. Our cohort was enriched for more severely affected individuals, but in the literature there were many individuals with missense variants in the motor domain with milder symptoms, more akin to pure HSP. It is likely these variants have an alternative mechanism of action, perhaps by hyperactivating motors, as was suggested in a recent study^63^.

While we observed some phenotype/genotype correlations, the correlations are imperfect, but this is not surprising. There are likely other contributors to the clinical phenotype including interacting genetic factors, prenatal and postnatal exposures, and the effects of therapeutic interventions. Our cohort included two sets of monozygotic twins, and in both instances, there were differences between the twins. In one family, one twin developed seizures in her teens while her sister did not develop seizures until her twenties. Many of these clinical features developed over time, and individuals in our cohort ranged in age in our cross-sectional study. With sufficient longitudinal data, we will assess how the scores and the disease change over time. An objective biomarker of neurodegeneration might provide a useful tool in conjunction with the clinical severity score.

The allelic heterogeneity of this condition is striking. We observed a range of severity across inheritance patterns, variant types, and variant locations. To assist with clearer communication about KAND, we propose three KAND subtypes: simple KAND, complex KAND, and HSAN KAND. We suggest distinguishing between individuals with only progressive lower extremity spasticity, ataxia, and peripheral neuropathy (simple KAND) and developmental delay, intellectual disability, seizures, optic nerve atrophy, and cerebellar atrophy (complex KAND). Complex KAND would include all individuals previously diagnosed with MRD9/NASCAV, PEHO, or *KIF1A*-related Rett syndrome. HSAN KAND would include individuals with variants in the alternatively spliced exon described previously. This new classification system could inform clinical management, and we suggest that all individuals with complex KAND would benefit from increased surveillance to detect the presence of seizures and progressive vision loss.

This study expands our understanding of the allelic spectrum and range of clinical phenotypes in KAND, and begins to address the molecular mechanisms of the disorder. With this broad phenotypic spectrum and the large number of variants, it is likely KAND is under recognized and under diagnosed. Better understanding of the phenotypic breadth and the disease subtypes could lead to improvements in diagnosis. As we move beyond diagnoses and work towards treatments, the ability to accurately predict prognosis and classify variants by molecular mechanism will facilitate the development of treatments.

## Data Availability

Data are available from the corresponding author upon reasonable request.

## Description of Supplemental Data

Supplemental data include 2 figures, 5 tables, and additional detailed methods.

## Acknowledgments

We thank KIF1A.org for their ongoing partnership and Autism Brain Net for the collection of postmortem brain specimens. We thank Scott Robinson, Jennifer Shahar, Emily Horowitz, and Rachel Cogny for their assistance with this project. We thank Kristen Verhey for her assistance with optimizing the neurite tip accumulation and in vitro gliding assays. Molecular graphics generated with UCSF Chimera, developed by the Resource for Biocomputing, Visualization, and Informatics at the University of California, San Francisco, with support from NIH P41-GM103311. Grant support includes TL1TR001875 (LB), R01N114636 (LR, AG, WKC), KIF1A.org, UL1TR001873. Research conducted at the Murdoch Children’s Research Institute was supported by the Victorian Government’s Operational Infrastructure Support Program.

## Web Resources

OMIM, http://www.omim.org.

